# Four doses of the inactivated SARS-CoV-2 vaccine redistribute humoral immune responses away from the Receptor Binding Domain

**DOI:** 10.1101/2022.02.19.22271215

**Authors:** Ji Wang, Caiguangxi Deng, Ming Liu, Yihao Liu, Liubing Li, Zhangping Huang, Liru Shang, Juan Jiang, Yongyong Li, Ruohui Mo, Hui Zhang, Min Liu, Sui Peng, Haipeng Xiao

## Abstract

A recent MMWR reported that the effectiveness of a 3^rd^ dose of SARS-CoV-2 mRNA vaccine waned quickly in the Omicron-predominant period. Similarly, a substantial decline of immune responses induced by a 3^rd^ dose of inactivated vaccines was also observed in our study. In response to the fast waning immune response and the great threat of Omicron variant of concern (VOC) to frontline healthcare workers (HCWs), 38 HCWs who were in our previous cohort investigating responses to the first three doses of inactivated vaccines participated in the current study and volunteered to receive a 4^th^ homologous booster. Here, we demonstrated that the 4^th^ dose is safe and capable of recalling waned immune responses 6 months after the 3^rd^ dose. However, a greater suppression on the induction of overall Neutralizing antibodies (NAbs) and NAbs targeting the receptor-binding domain (RBD) was found in participants with stronger immune responses after the 3^rd^ dose. As a result, a stepwise elevation of RBD-NAbs from the 1^st^ to the 3^rd^ vaccination achieved a “turning point”. The peak RBD-NAbs level induced by the 4^th^ dose was inferior to the peak of the 3^rd^ dose. Accompanied with reduced induction of RBD-NAbs, the immune system shifted responses to the nucleocapsid protein (NP) and the N-terminal domain (NTD) of the spike protein. Although NTD directed antibodies are capable of neutralization, they only compensated the loss of RBD-NAbs to ancestral SARS-CoV-2 virus but not to the Omicron variant due to a substantial conformational change of Omicron NTD. This longitudinal clinical study monitored the immune response of the same cohort for every doses, shaping a relationship between the trajectory of immune focus and the dynamics of the neutralizing potency against the evolving virus. Our data reveal that immune responses could not be endlessly elevated, while suppression of heightened immune responses focusing on one subunit together with a shift of immune responses to other subunits would occur after repeated vaccination. Thus, an updated vaccine with more diverse epitopes capable of inducing NAbs against VOCs would be a future direction for boosters.

## Introduction

Vaccination is one of the most cost-effective ways to prevent infectious diseases, including COVID-19. Billions of vaccine doses have been distributed worldwide and showed promising effectiveness against SARS-CoV-2 infection and related hospitalization. However, the vaccine-induced immune responses waned rapidly after receiving two doses of mRNA vaccines ^1^. Our previous study also showed humoral immune responses elicited by inactivated SARS-CoV-2 vaccines declined quickly within 6 months after a standard two-dose vaccination regimen ^2^. In addition to the fading immune response, frequent emerging of mutated SARS-CoV-2 viruses, especially those variants of concern (VOC), further challenges the vaccination system based on the ancestral viral strain ^3,4^.

Therefore, a booster or a 3^rd^ dose of vaccines were provided globally. Our studies and others have demonstrated that the 3^rd^ dose elevated both humoral and cellular immune responses to a much greater level from the two-dose regimen, equipping the population with potent protection not only for ancestral virus but also for VOCs, such as Delta ^2,5^. Unfortunately, recently emerged VOC Omicron carries more than 30 mutations, rendering an overwhelming capability of escaping immune responses established by vaccination or natural infection ^4^. Numerous breakthrough infections have been reported worldwide ^6^. While antibodies induced by the 3^rd^ dose of vaccines do neutralize Omicron to some extent and T cell responses are cross-reactive, preliminary data have shown that the protection provided by the booster were not complete and also waned at a fast pace ^7-9^. A recent report from US CDC revealed that the vaccine efficiency against emergency department and urgent care encounters in people who had received 3 doses of mRNA vaccines declined from 87% to 66% within 4 months, and further dropped to 31% after 5 months in the Omicron-predominant period ^7^. Thus, in early January, Israel began to provide a 4^th^ dose of vaccines to the most vulnerable populations, including Healthcare workers (HCWs) ^10,11^.

In this study, we continued to monitor our established cohort compose of frontline HCWs for the immune responses induced by a 3^rd^ dose of inactivated SARS-CoV-2 vaccine. Given that only 15% humoral immune responses remained after 6 months and a great threat of Omicron to HCWs, a 4^th^ dose of inactivated vaccines was subsequently provided. The immune responses against both ancestral SARS-CoV-2 strain (Wuhan-Hu-1 reference strain) and Omicron variant were monitored along with a longitudinal assessment of humoral responses to multiple antigens and domains.

## Results

### The immune response induced by the 3^**rd**^dose of inactivated vaccine waned rapidly

We have previously conducted a non-randomized trial and recruited HCWs from a prospective cohort. They received a primary 2-dose series of the inactivated SARS-CoV-2 vaccine (BBIBP-CorV, Sinopharm, Beijing) and a 3^rd^ dose booster of the same vaccine 5 months later ^2,12^. In this study, we first monitored the longevity of immune responses after the 3^rd^ dose. Thirty-eight HCWs from the previous cohort volunteered in the current non-randomized trial. Serum neutralizing antibodies (NAbs) against an ancestral SARS-CoV-2 viral strain (Wuhan-Hu-1), named wildtype (WT) hereafter, or the Omicron variant were measured by a pseudovirus assay and quantified as half pseudovirus neutralization titers (PVNT50) ^13^. The geometric mean of neutralization titers (GMNT) against WT drastically decreased by 85% 26 weeks (wks) after the 3^rd^ dose, as compared to GMNT at 2 wks after the 3^rd^ dose (**Fig. 1a, blue circles in 1**^**st**^ **and 2**^**nd**^ **panels**). As expected, GMNT against Omicron was much lower than that against WT at these two time points, though the immune response against Omicron seemed to dropped at a slower pace (**Fig. 1a, orange triangles in 1**^**st**^ **and 2**^**nd**^ **panels**). GMNT against Omicron decreased by 53% within 6 months. No significant difference was observed between male and female HCWs at any time point (**Supplementary Fig. 1a and b**).

**Figure 1.**
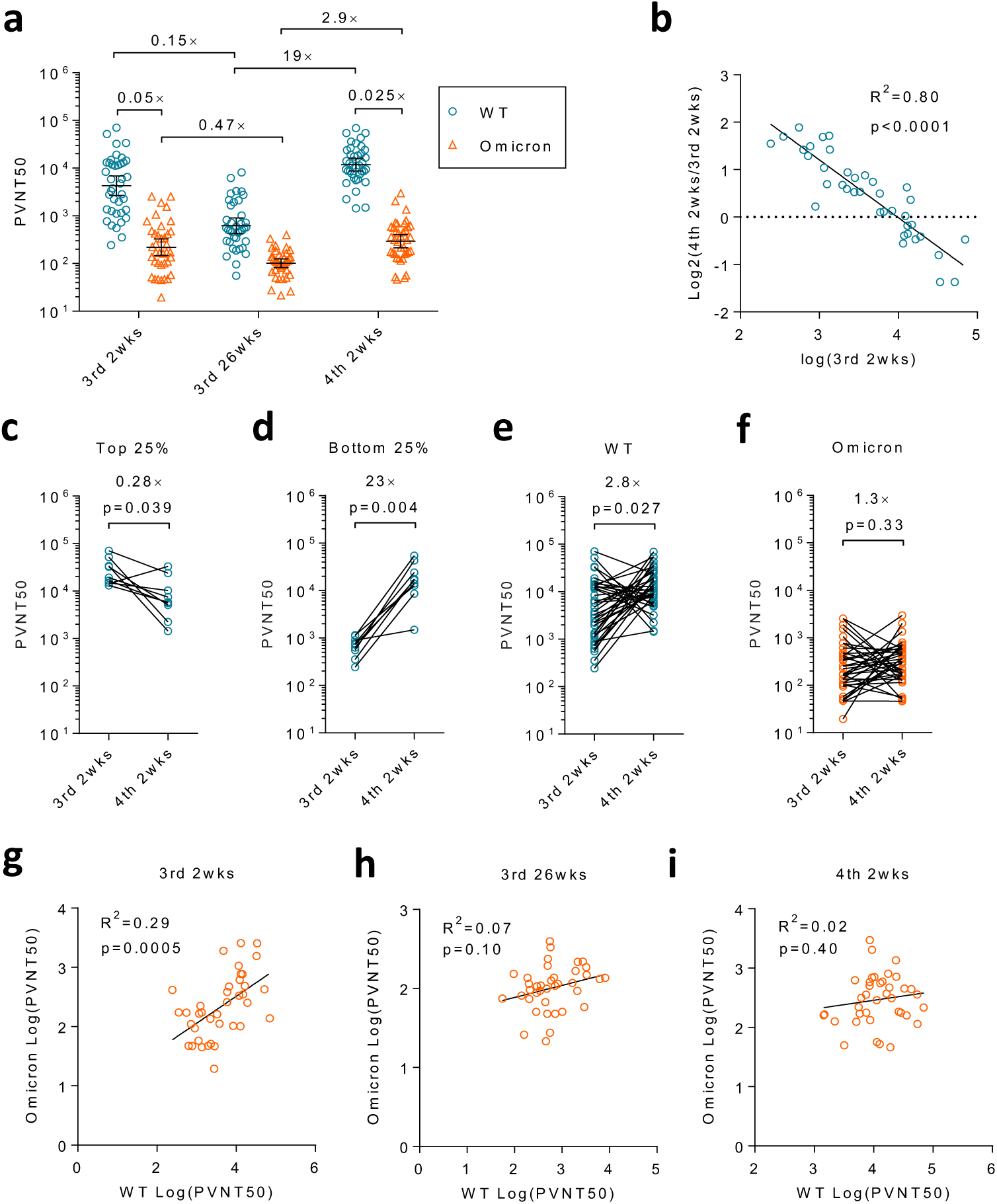
A 4^th^ dose of inactivated SARS-CoV-2 vaccine boosted NAbs against wildtype (WT) virus but not Omicron. Thirty-eight HCWs who have already received 3 doses of inactivated SARS-CoV-2 vaccine volunteered in this clinical study and received a 4^th^ homologous dose 6 months (26 wks) after the 3^rd^ dose. (**a**) Neutralization assays were performed to measure NAbs titers against pseudoviruses with S protein from a WT strain (blue circle) or Omicron variant (orange triangle). (**b**) Linear regression was performed on the fold-change of PVNT50 from 3^rd^ 2wks to 4^th^ 2wks and PVNT50 at 3^rd^ 2wks. (**c**) The change of NAbs was shown for participants with the top 25% highest PVNT50 against WT virus. (**d**) The change of NAbs was shown for participants with the lowest (bottom 25%) PVNT50 against WT virus. NAbs at 3^rd^ 2wks and 4^th^ 2wks against WT virus (**e**) or Omicron variant (**f**) were compared respectively. A series of Linear regression was performed between PVNT50 to WT or Omicron at 3^rd^ 2wks (**g**), 3^rd^ 26wks (**h**), and 4^th^ 2wks (**i**). Data were shown as Geometric mean ± 95% confidence level (Cl). Wilcoxon matched-pairs signed rank test was used for (**c-f**).

### A 4^**th**^dose of vaccination recalled the waned immune response

Since Omicron had been threatening the healthcare systems by the end of the previous trial and the waned immune responses were unlikely to provide sufficient protection against infection to HCWs, especially in the context of Omicron, the same cohort of 38 HCWs volunteered in a new non-randomized trial (ChiCTR2200055564) to investigate the potential benefit of a 4^th^ dose. No severe side effects related to vaccination were recorded during the trial (**Table 1**). The 4^th^ dose robustly recalled NAbs titers against WT by 19 folds (**Fig. 1a, blue circles in 3^rd^ panel**). Cross-reactive NAbs to Omicron were successfully also recalled despite to a lesser extent, 2.9 folds (**Fig. 1a, orange triangles in 3**^**rd**^ **panel**).

**Table 1.** Demographics and vaccination-related adverse events

|  |  | Total |
| --- | --- | --- |
| Participants, N |  | 38 |
| Age, mean (SD) |  | 27.63(6.70) |
| Gender, male (%) |  | 18 (47.4) |
| Adverse events | N (%) | 7(18.4) |
| Injection site symptoms | Pruritus, N (%) | 2(5.3) |
|  | Swollen, N (%) | 1(2.6) |
| Systematic symptoms | Rash, N (%) | 1(2.6) |
|  | Dizziness, N (%) | 2(5.3) |
|  | Nausea, N (%) | 1(2.6) |
|  | Fatigue, N (%) | 1(2.6) |

### Immune responses induced by the 3rd dose negatively affected the outcome of the 4th dose

As SARS-CoV-2 continuously circulates globally waves by waves, multiple vaccinations are needed. Thus, it is of great importance to know the impact of previous vaccination on the following booster. We first analyzed the correlation between NAb titers at various time points, finding that neither titers at 3^rd^ 2wks nor 3^rd^ 26wks correlated with titers at 4^th^ 2wks (**Supplementary Fig. 2a and b**). Surprisingly, a reverse correlation between NAb titers at 2wks after the 3^rd^ dose (3^rd^ 2wks) and the fold change from the 3^rd^ peak to the 4^th^ peak (4^th^ 2wks/3^rd^ 2wks) was observed (R^2^=0.80, p<0.0001), indicating whether or not the peak humoral response of the 4^th^ vaccination could be further elevated depended on the peak of the previous vaccination (**Fig. 1b**). In contrast, residual immune responses at 3^rd^ 26wks, the immune responses right before the 4^th^ vaccination, had little impact on the outcome of the 4^th^ dose (**Supplementary Fig. 2c**). Collectively, these data demonstrated that it is the peak response after the 3^rd^ vaccination that determined whether a breakthrough from the previous peak would take place after the 4^th^ dose.

As a result of this negative correlation, inferior titers at 4^th^ 2wks to those at 3^rd^ 2wks were always detected among the top 25% participants with the highest NAb titers at 3^rd^ 2wks (**Fig. 1c**). Conversely, the bottom 25% participants who had a low immune response benefited most from a drastic increase in NAb titers after the 4^th^ vaccination (**Fig. 1d**). These results suggest that individuals who respond less well to the first 3 doses are preferable for the 4^th^ dose. The indicator is the peak response after the 3^rd^ dose, whose cut-off value is a PVNT50 of 1×10^4^ in our pseudovirus system (**Fig. 1b**).

### The 4^th^dose had distinct effects on WT virus and Omicron variant

As mentioned above, the elevation from 3^rd^ 26wks to 4^th^ 2wks was much greater for WT than that for Omicron (19-fold vs 2.9-fold) (**Fig. 1a**). We next compared peak NAb titers after the 3^rd^ dose and the 4^th^ dose. GMNT increased by 2.8-fold for WT, whereas no significant change was observed for Omicron (**Fig. 1e and f**). This data indicated that whilst the 4^th^ vaccination further enhanced WT-specific immune responses, the cross-neutralizing immune response was not equally strengthened. As a result, the GMNT ratio between two viral strains dropped from 0.05 to 0.025 after the 4^th^ dose (**Fig. 1a, 1**^**st**^ **and 3**^**rd**^ **panels**). To further validate the different effects of the 4^th^ dose on two strains, linear regression analysis was performed between NAb titers against WT and Omicron at various time points. As expected, a significant albeit moderate correlation (p=0.0005) was seen at 3^rd^ 2wks, but gradually lost over time and was disrupted (p=0.40) after the 4^th^ dose (**Fig. 1g-i**). Taken together, these results suggested a shift of immune responses from shared epitopes to non-cross-reactive ones after the 4^th^ vaccination.

### Repeated vaccination of inactivated vaccines shifted humoral immune responses to Nucleocapsids

The inactivated SARS-CoV-2 vaccine contains all viral structure proteins, among which Spike protein (S), Nucleocapsid (NP), Envelope (E) are immunogenic as revealed by our previous study ^2^. Among these immunogenic viral proteins, only S protein plays the pivotal role in inducing NAbs, while the contribution of other proteins may be minimal ^14^. Thus, we next investigated whether the 4^th^ dose altered the distribution of humoral immune responses to various antigens. As expected, anti-S and anti-NP antibody titers decreased in a large portion of participants between 2 and 26 weeks after the 3^rd^ dose (**Fig. 2a, 1**^**st**^ **and 2**^**nd**^ **bars**), except for anti-E antibody titers which increased in >50% participants in this period (**Fig. 2a, 3**^**rd**^ **bar**). Unexpectedly, the 4^th^ dose did not effectively recall anti-S and anti-E antibodies in a majority of participants, while it did successfully recall anti-NP antibodies in >50% individuals of the same population (**Fig. 2b, 2**^**nd**^ **bar**).

**Figure 2.**
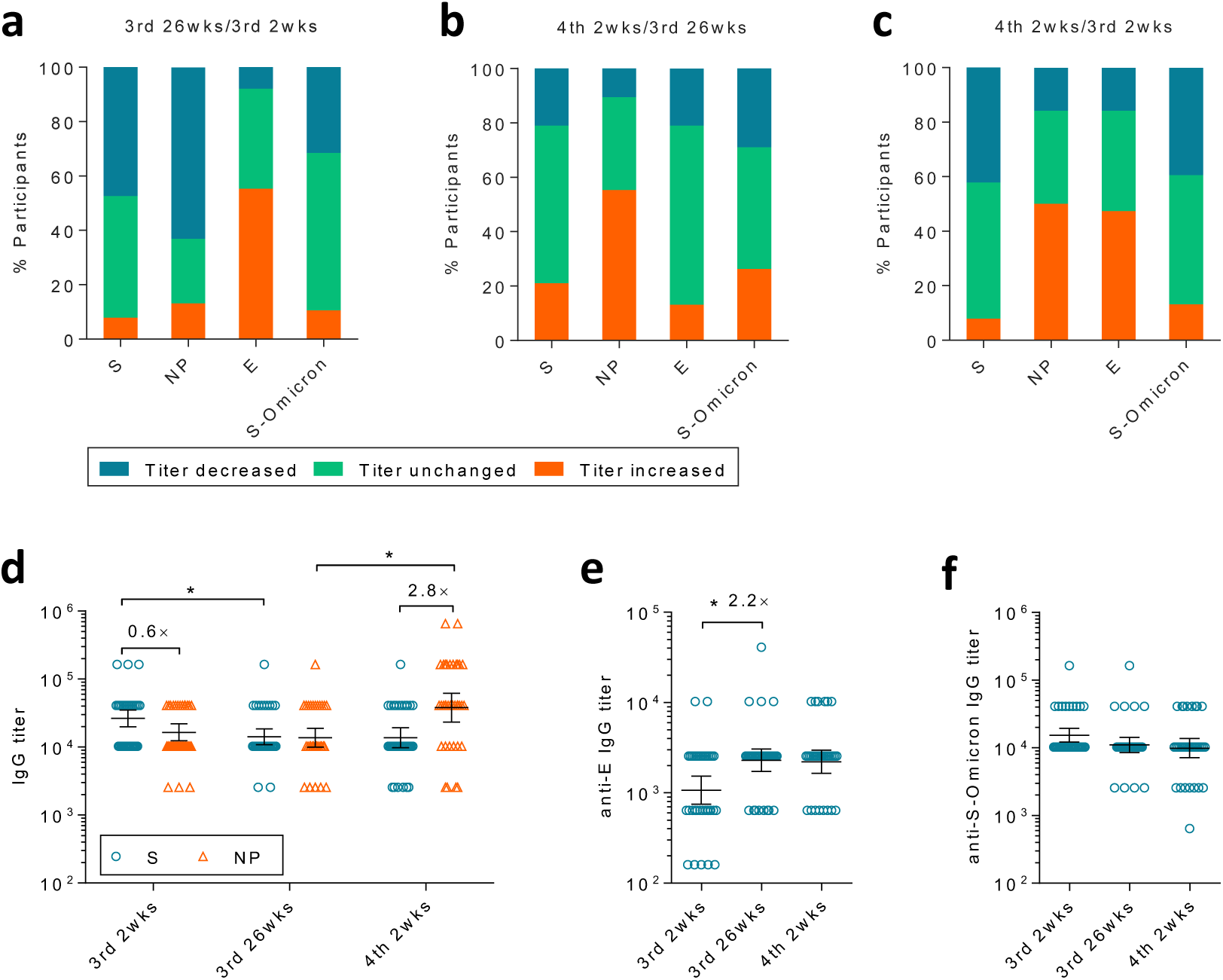
Humoral immune responses shifted from S protein to NP. Anti-S, NP, E and S-Omicron antibodies were measured by ELISA at 2 weeks after the 3^rd^ dose (3^rd^ 2wks), 26 weeks after the 3^rd^ dose (3^rd^ 26wks) or 2 weeks after the 4^th^ dose (4^th^ 2wks). (**a-c**) Antibody titers for each protein were compared between 3^rd^ 2wks and 26wks (**a**), 4^th^ 2wks and 3^rd^ 26wks (**b**), 4^th^ 2wks and 3^rd^ 2wks (**c**), respectively. Percentage of Participants with increased titers (former/later >1, orange), decreased titers (former/later <1, blue) or unchanged titers (former/later =1, green) were summarized. (**d**) Anti-S (blue circle) and anti-NP (orange triangle) antibody titers were compared between each time point. (**e**) Anti-E antibody titers at each time point were compared. (**f**) Antibody titers for S-Omicron. Data were shown as Geometric mean ± 95% Cl. Friedman test followed by Dunn’s multiple comparisons test was used for d-f. * p<0.05.

As a result, most participants had inferior peak anti-S antibody titers after the 4^th^ dose as compared to peak titers after the 3^rd^ dose (**Fig. 2c, 1**^**st**^ **bar**), in sharp contrast to anti-NP antibodies whose peak titers were further elevated by the 4^th^ dose in >50% of individuals (**Fig. 2c, 2**^**nd**^ **bar**). Anti-S antibody titers significantly dropped from 2wks to 26wks after the 3^rd^ dose, but did not raised after the 4^th^ dose (**Fig. 2d, blue circles**). In contrast, anti-NP antibody titers only dropped slightly before the 4^th^ dose but greatly raised thereafter (**Fig. 2d, orange triangles**). As a result, the ratio of peak antibody titers between anti-NP and anti-S was reversed by the 4^th^ dose. Anti-NP was 40% lower than anti-S right after the 3^rd^ dose, but became 2.8-fold higher after the 4^th^ dose (**Fig. 2d, 1**^**st**^ **and 3**^**rd**^ **panels**).

Antibodies against E, another immunogenic antigen in the vaccine, showed an increasing trend from the 3^rd^ to the 4^th^ dose (**Fig. 2e**). However, the impact of this change on anti-S antibodies and the protection remained elusive since anti-E antibody titers were one magnitude lower than that of anti-S or anti-NP antibodies (**Fig. 2d and e**). We further measured cross-reactive antibodies to Omicron Spike protein (S-Omicron), observing a similar decreasing trend as ancestral S protein albeit titers were slightly lower (**Fig. 2a-c, f**). Distinct trends of anti-NP and anti-S titers suggested that the immune system began to focus on a more immunodominant antigen, such as NP, after repeated vaccination of the inactivated vaccines that comprise multiple viral proteins. Unfortunately, humoral immune responses against NP, a protein hiding inside the virus and infected cells, are less likely to mediate protection ^14^.

### The 4^**th**^dose redistributed the humoral immune response away from the receptor-binding domain (RBD)

Reduced efficacy of the 4^th^ dose on elevating anti-S antibodies partially explained why the 4^th^ did not dramatically elevate the peak of NAbs as the 3^rd^ dose did ^2^, but did not explain why the NAbs to WT still increased slightly while those to Omicron did not (**Fig. 1e and f**). To explore this aspect, we further investigated humoral responses to various domains on S, including S1 and S2. The S1 could be further divided into the N-terminal domain (NTD) and RBD. As expected, antibody titers against these domains decreased in most participants after 3^rd^ dose (**Fig. 3a**). Anti-S1 and anti-S2 antibodies showed similar trends compared to anti-S antibodies, and no significant bias was observed for these two domains (**Fig. 3a-d**). However, the 4^th^ dose had different effects on antibodies to sub-domains in S1. In detail, anti-NTD titers increased in >90% of participants (**Fig. 3b, 3**^**rd**^ **bar**), whereas anti-RBD titers only increased in <40% of individuals (**Fig. 3b, 4**^**th**^ **bar**). The overall effect is that ∼50% of participants had increased anti-NTD titers while others remained unchanged between the 3^rd^ peak and the 4^th^ peak (**Fig. 3c, 3**^**rd**^ **bar**). In sharp contrast, >50% of participants exhibited a reduced anti-RBD titer (**Fig. 3c, 4**^**th**^ **bar**). Both geometric mean titers of anti-RBD and anti-NTD IgG significantly dropped at 3^rd^ 26wks, but were boosted by the 4^th^ dose at different slopes. Anti-NTD titers increased by 8-fold after the 4^th^ dose, whereas the increase for anti-RBD was only 1.9-fold (**Fig. 3e, 2**^**nd**^ **and 3**^**rd**^ **panels**). Because of such a huge difference in response to the 4^th^ dose, the ratio of anti-NTD/anti-RBD vigorously increased from 0.2 to 1.4 (**Fig. 3e, 1**^**st**^ **and 3**^**rd**^ **panels**). In another word, while anti-NTD titers were much lower than anti-RBD titers after the 3^rd^ dose, they were boosted to a comparable or even slightly higher level over anti-RBD after receiving the 4^th^ dose. The geometric mean titer of anti-NTD at 4^th^ 2wks was higher than that at 3^rd^ 2wks (**Fig. 3e, orange triangles**). In contrast, anti-RBD titers declined from 3^rd^ 2wks to 4^th^ 2wks (**Fig. 3e, blue circles**). Change of cross-reactive antibodies to RBD-Omicron after the 3^rd^ and the 4^th^ dose showed a similar trend as those to WT RBD (orange dashed line) albeit titers of anti-RBD-Omicron were lower at all time points (**Fig. 2f**).

**Figure 3.**
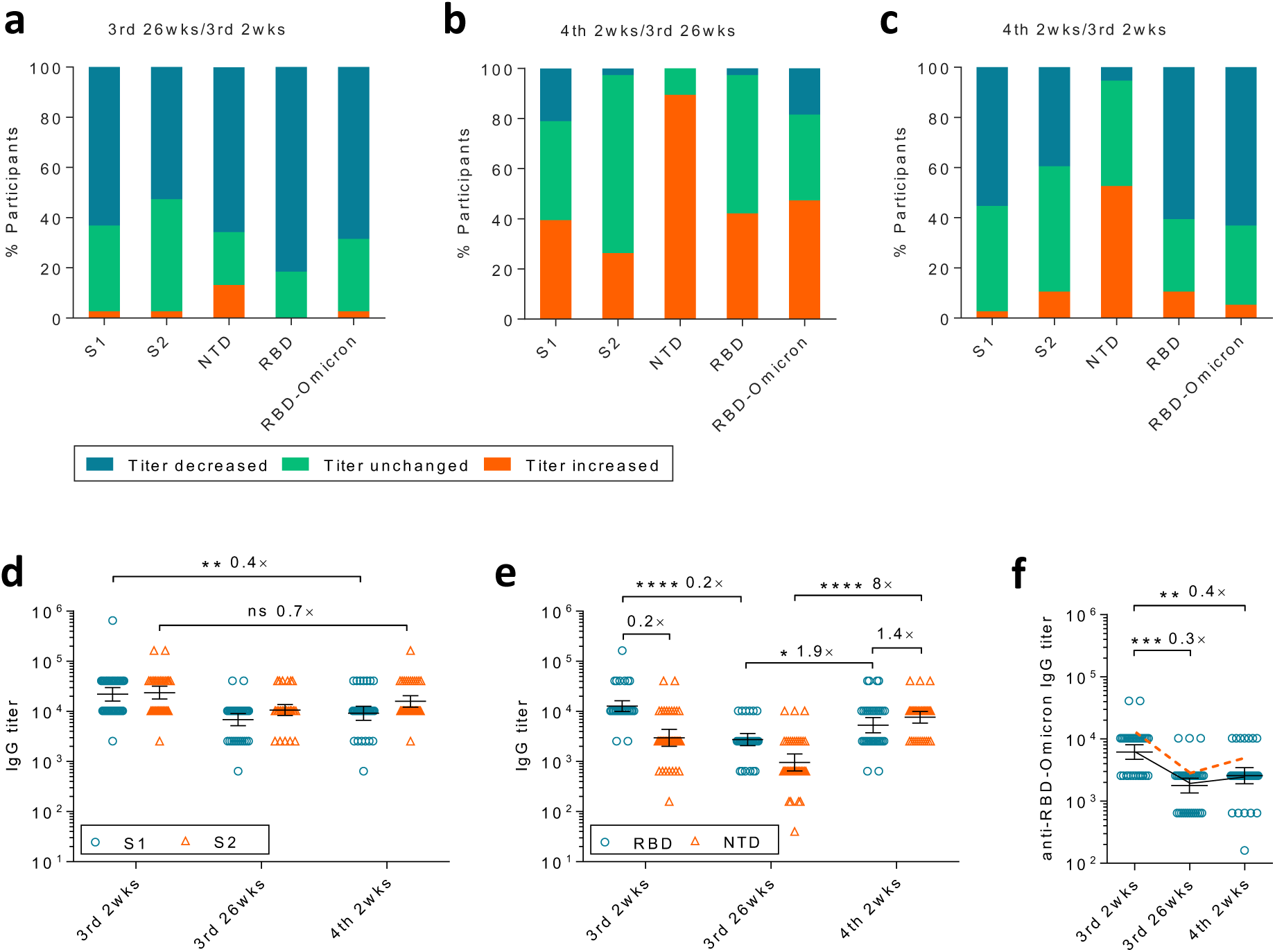
The S protein directed antibody responses shifted from RBD to NTD. Anti-S1, S2, NTD, RBD and RBD-Omicron antibodies were measured by ELISA at 3^rd^ 2wks, 3^rd^ 26wks or 4^th^ 2wks. (**a-c**) Antibody titers for each protein were compared between 3^rd^ 2wks and 26wks, 4^th^ 2wks and 3^rd^ 26wks, 4^th^ 2wks and 3^rd^ 2wks, respectively. Percentage of participants with increased titers (former/later >1, orange), decreased titers (former/later <1, blue) or unchanged titers (former/later =1, green) were shown. (**d**) Anti-S1 (blue cricle) and anti-S2 (orange triangle) antibody titers were compared between each time point. (**e**) Anti-RBD (blue circle) and anti-NTD (orange triangle) antibody titers at each time point were compared. (**d**) Antibody titers for S-Omicron. The orange dashed line represents the kinetics of the geometric mean titer of anti-S IgG. Data were shown as Geometric mean ± 95% Cl. Friedman test followed by Dunn’s multiple comparisons test was used for d-f. * p<0.05, **p<0.01, ***p<0.001, ****p<0.0001. ns, not significant.

For WT virus, NAbs could be induced by both NTD and RBD. However, recent studies have demonstrated that most cross-neutralizing antibodies against Omicron target majorly to RBD rather than other domains in S, such as NTD ^4^. A biased increment on anti-NTD rather than anti-RBD antibodies after the 4^th^ dose was in line with our neutralizing data that the 4^th^ dose profoundly enhanced neutralization to WT virus but not Omicron.

### Robust immune responses elicited by the 3rd dose attenuated induction of RBD-NAbs after the 4th dose

In addition to binding antibodies to RBD, we further investigated whether NAbs to RBD also decreased. Thus, a one-step competitive Chemiluminescent immunoassay was used to detect the concentration of NAbs that compete with ACE2 binding to RBD ^15^. Whilst the 3^rd^ dose induced robust immune responses at 2 and 4 weeks after immunization, the RBD-NAbs waned quickly (**Fig. 4a**). An average drop of 60% from the peak was observed within 3 months after the 3^rd^ dose (**Fig. 4b**). Although only 25% of immune responses remained after 6 months, residual immune responses were still significantly higher than that at 5 months after the 2^nd^ dose (**Supplementary Fig. 3**). The 4^th^ dose effectively elevated RBD-NAbs in all vaccinees, whose geometric mean RBD-NAbs level increased by 2.7-fold within 2 weeks after immunization as compared to those at 6 months after the 3^rd^ dose (**Fig. 4b**). The 4^th^ dose was equally effective for both genders in inducing RBD-NAbs (**Supplementary Fig. 4**). However, the immune responses induced by the 4^th^ dose seemed to wane at a faster pace than that induced by the 3^rd^ dose. Geometric mean RBD-NAbs remained comparable between 3^rd^ 2wks and 3^rd^ 4wks, but a significant decrease was found unexpectedly at 4^th^ 4wks (**Fig. 4b**).

**Figure 4.**
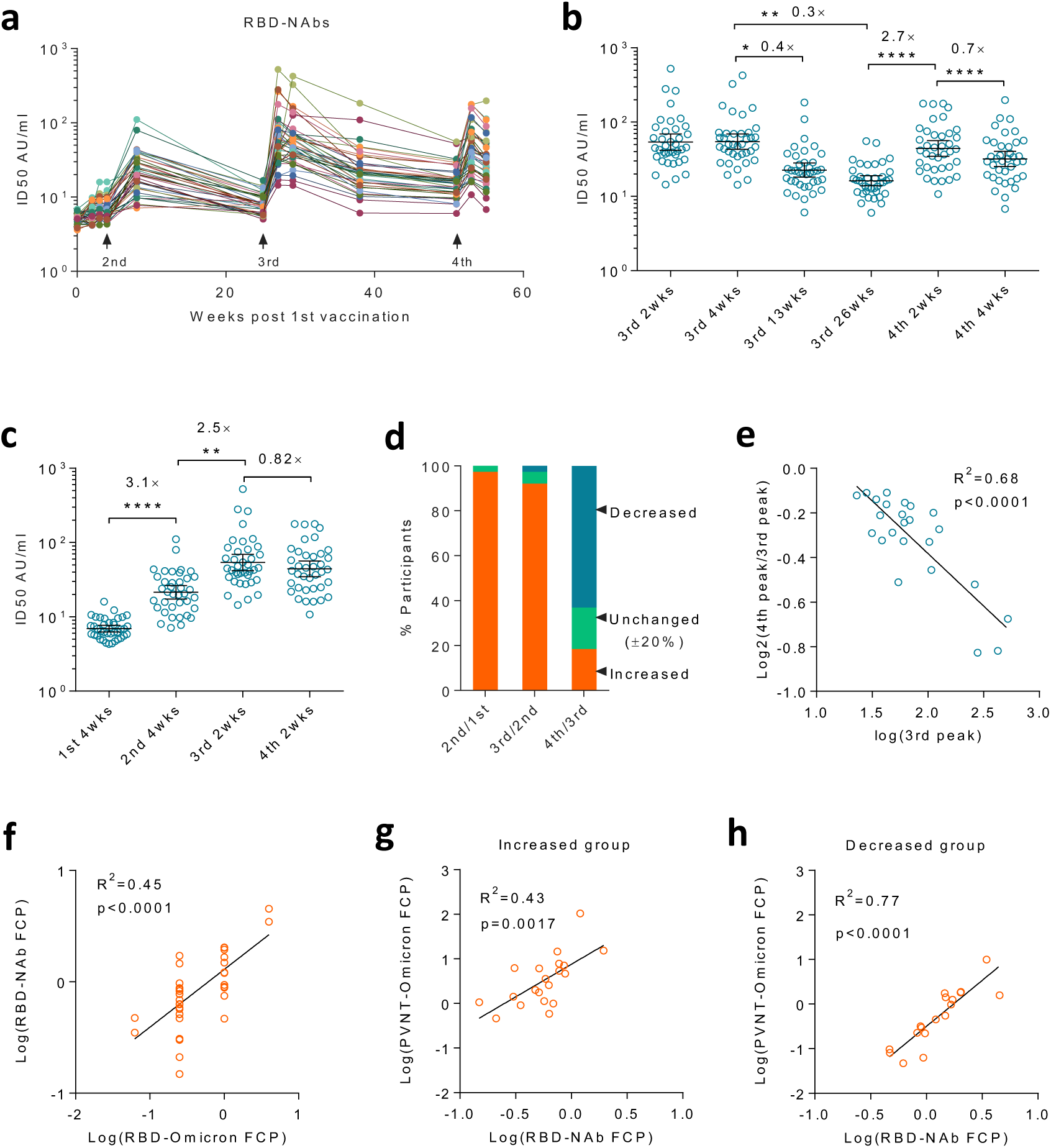
Potent immune responses elicited by the 3rd dose suppressed the induction of RBD-NAbs after the 4th dose. **(a)**NAbs to RBD (RBD-NAbs) were measured by a one-step competitive Chemiluminescent immunoassay for sera collected at various time points after 1^st^, 2^nd^, 3^rd^, and 4^th^ doses. The kinetic of RBD-NAbs was shown for each participant. (**b**) RBD-NAbs were compared among various time points, including 3^rd^ 2wks, 3^rd^ 4wks, 3^rd^ 13wks, 3^rd^ 26wks, 4^th^ 2wks, and 4^th^ 4wks. (**c**) Peak values of RBD-NAbs after each vaccination were compared. (**d**) Percentage of participants with increased peak value (increased >20%, orange), decreased value (decreased >20%, blue), or unchanged (±20%, green) as compared to the previous peak were summarized. (**e**) Linear regression was performed between the peak value after the 3^rd^ dose and the fold change of peak value from the 3^rd^ to the 4^th^ dose in the decreased group shown in (**d**). (**f**) Correlation between fold-change of peak values (FCP) of RBD-NAbs and FCP of anti-RBD-Omicron IgG was analyzed by linear regression. (**g, h**) Participants were separated into an increased group or a decreased group, defined as PVNT-Omicron FCP increased or decreased compared to the RBD-NAb FCP respectively, taking PVNT-Omicron FCP/ RBD-NAb FCP >2 as the threshold. Linear regression was performed between FCP of PVNT50 to Omicron and FCP of RBD-NAb in each group. Data were shown as Geometric mean ± 95% Cl. RM one-way ANOVA was used for b and c. * p<0.05, **p<0.01, ***p<0.001, ****p<0.0001. ns, not significant.

Whilst the first three doses resulted in a stepwise elevation of peak RBD-NAbs, the 4^th^ dose did not. The 2^nd^ and 3^rd^ dose enhanced RBD-NAbs by 3.1-fold and 2.5-fold to the previous peak, respectively (**Fig. 4c**). However, the 4^th^ dose did not further elevate the peak value of RBD-NAbs, but an 18% decrease was found instead (**Fig. 4c**). The inferior peak value after the 4^th^ dose did not happen sporadically. In 63% (24/38) of participants, the peak RBD-NAbs after the 4^th^ dose was 20% lower than that after the 3^rd^ dose, whereas only 18% (7/38) increased by >20% (**Fig. 4d, 3**^**rd**^ **bar**). In sharp contrast, >90% of participants (37/38 and 35/38) benefited from each booster shot of the 2^nd^ and the 3^rd^ dose (**Fig. 4d, 1**^**st**^ **and 2**^**nd**^ **bars**).

As indicated by our pseudovirus neutralization data, immune responses induced by the previous vaccination may negatively affect NAbs induced by the following dose (**Fig. 1b and c**). However, neutralization assay involved antibodies induced by multiple antigens or domains. Here, we explored whether it is true for the immune response to a single domain as well. A significant negative correlation between fold-change of peak values and the values of previous peaks was observed after the 4^th^ dose in participants with decreased peak RBD-NAbs, indicating that potent immune responses elicited by the 3^rd^ dose did suppress the induction of RBD-NAbs after the 4^th^ dose (**Fig. 4e**).

### NAbs to RBD mediated cross-neutralization against Omicron

Our pseudovirus neutralization assay and ELISA suggested that the distinct effect of the 4^th^ dose on WT virus and Omicron variant depended on antibodies to RBD. A reduced peak RBD-NAbs after the 4^th^ dose coincided with pseudovirus neutralization and ELISA results. Lastly, the evidence for the contribution of RBD-NAbs was investigated. A series of linear regression was performed on the fold change of peak values (FCP) between 3^rd^ and 4^th^ doses to eliminate the effect of other factors, such as the contribution of NAbs outside RBD. A positive correlation was found between RBD-NAbs FCP and FCP of RBD-Omicron binding antibodies, enabling us to further investigate the relationship between NAbs against Omicron in pseudovirus neutralization assay (PVNT-Omicron) and RBD-NAbs (**Fig. 4f**). There was no significant correlation between FCPs of PVNT-Omicron and RBD-NAbs when taking the population as a whole, but strong correlations were observed in the increased group (R^2^=0.43, p=0.0017) and the decreased group (R^2^=0.77, p<0.0001) respectively (**Fig. 4g and h**). Two groups were defined as PVNT-Omicron FCP increased or decreased compared to the RBD-NAb FCP respectively, taking PVNT-Omicron FCP/ RBD-NAb FCP >2 as the threshold. Two different models of the action were likely involved in these groups. Nevertheless, these data revealed that RBD-NAbs contributed to the cross-neutralization against Omicron in immune responses induced by inactivated SARS-CoV-2 vaccines.

## Discussion

In this study, we investigated the safety and effectiveness of the 4^th^ dose of inactivated SARS-CoV-2 vaccine in HCW volunteers when a great loss of protective humoral immune responses was found for both ancestral SARS-CoV-2 and Omicron variant 6 months after the 3^rd^ dose. At the moment, Omicron is continuously threatening the healthcare system in which HCWs are the most vulnerable population. Our results demonstrated that the inactivated SARS-CoV-2 vaccine had an acceptable safety profile that no severe side effect was found after the 4^th^ dose despite a higher onset of adverse events (18.4%) occurring as compared to the 3^rd^ dose (12%). When taking the immunity 6 months after the 3^rd^ dose as the benchmark, the 4^th^ dose is meaningful. It successfully recalled the waned immune responses against both ancestral virus and Omicron variant with a low cost of safety. However, when taking the four doses of vaccination together, several implications on immunology and vaccinology are provided by the current study.

It is a consensus that immune responses could not be endlessly boosted. A plateau or even a “turning point” would occur after repeated vaccination, but such a phenomenon has been rarely evidenced by a well-designed clinical study involving multiple administration of the same vaccine without any interference of pre-existing immunity and asymptomatic infections during the study. Many studies as well as ours all revealed a stepwise elevation of peak immune responses from 1^st^ to 3^rd^ dose of SARS-CoV-2 vaccines ^2,16^. No sign of a plateau or a turning point was observed before the current study enrolling a 4^th^ dose. Our data indicated that the 3^rd^ dose is the “turning point” for repeated vaccination of inactivated SARS-CoV-2 vaccines made from the ancestral viral strain. We observed a clear suppression of humoral response to the 4^th^ dose by a heightened immune response after the 3^rd^ dose, confirmed by two different assays— pseudovirus neutralization assay and RBD-NAbs assay (**Fig. 1b and Fig. 4e**). As the result of such suppression, peak levels of S-binding, RBD-binding and RBD-NAbs were all inferior to their counterparts after the 3^rd^ dose (**Fig. 2d, Fig. 3e and Fig. 4d**).

A recent study revealed that induction of hemagglutination inhibition (HI) antibodies, similar to RBD-NAbs in SARS-CoV-2, by a second homologous dose of influenza vaccines was attenuated by the previous dose ^17^. However, the small group size of participants receiving two doses in that study (n=3-8) and unknown history of natural infections prevented the study from establishing a reliable quantitative correlation. In contrast, our study enrolled a relatively large number of participants (n=38) who were tested for SARS-CoV-2 weekly and had never been infected before or during the study. Thus, pre-existing immunity or occasional infection during the study which is always a concern for clinical influenza vaccine studies, would not perturb the results of this study. Taking the advantage of acceptable group size and clear immunological background, we are able to draw a clear picture that humoral immune responses to a certain region of antigens, such as RBD, were elevated dose by dose till the maximal capacity is achieved (**Fig. 4c**). After that, immune responses were down-regulated by a ratio tightly associate with the maximal response induced by the previous vaccination (**Fig.1b and Fig. 4e**). The timing for the plateau may vary depending on the nature of antigens and adjuvants. For inactivated SARS-CoV-2 vaccine used in this study, the turning point is 3^rd^ dose for most participants. Interestingly, we found the PVNT50 of 1×10^4^ is a critical point indicating whether the peak of immune responses could be further elevated by the 4^th^ dose. Despite most vaccinees experiencing suppression, for those participants with a poor response to the 3^rd^ dose, the 4^th^ dose was still very effective (**Fig. 1d**). Of note, this cut-off value may vary from lab to lab and depend on the assay used.

Mechanisms underlying the down-regulation of immune responses are unclear yet. Since our data have shown that it was the peak NAb level after the 3^rd^ dose rather than the NAb level right before the 4^th^ dose determined the depth of the down-regulation, we speculate that atypical memory B cells or B cell exhaustion which is always induced by repeated antigen exposure during chronic viral infection may contribute majorly, rather than other mechanisms such as epitope masking ^18,19^.

Recent studies, as well as our unpublished data, revealed that a 3^rd^ dose of inactivated vaccine or mRNA vaccine could induce a higher level of cross-neutralizing antibodies against Omicron ^8,20^. Strikingly, data in the current study indicated that neutralization breadth was not further increased by the 4^th^ dose, but even narrowed (**Fig. 1a, g and i**). Down-regulation of overheated immune responses against one domain/epitope leaves room for inducing immune responses to other epitopes, facilitating the immune system to establish a more diverse immunity which is always beneficial. However, it is not the case for the induction of cross-NAbs against Omicron by inactivated SARS-CoV-2 vaccines composed of a full-length S protein and other structural viral proteins. An increase of humoral immune responses to NP protein and NTD domain was observed after the 4^th^ dose accompanied with down-regulation of humoral response to RBD (**Fig. 2d and Fig. 3e**). Unfortunately, increased humoral immune response to NP is less likely to be protective since access of antibodies to NP is prohibited by viral or cell membranes ^14^. On the other hand, whilst a large number of NTD-directed antibodies do neutralize WT virus, few numbers of such antibodies could cross-neutralize Omicron variant since mutations induced a substantial conformational change in NTD antigenic supersite which is the target of most NTD-directed neutralizing antibodies ^21^. Conversely, some RBD-induced antibodies still have cross-neutralizing capabilities which are also revealed by our analysis (**Fig. 4g and h**) ^4,16^. Therefore, upregulation of NTD-induced antibodies compensated the loss of RBD-directed neutralizing activity for WT virus but not for Omicron (**Fig. 1a, g and i**).

While these results were obtained from repeated vaccination of whole inactivated SARS-CoV-2 vaccines, it will be interesting and important to know, whether down-regulation of RBD-NAbs would occur in other types of vaccines comprising RBD, and shifting of humoral response to other domains would also happen in vaccines comprising the whole sequence of S protein, such as mRNA-1273 and BNT162b. It is important to note, however, that widely used cross-sectional cohorts are less capable of characterizing the dynamics of neutralizing breadth and humoral responses to multiple epitopes. Instead, a longitudinal cohort, such as the cohort used in the current study, is preferred.

Nevertheless, a very recent report on the 4^th^ dose of mRNA vaccines supported our findings. The study demonstrated that a 4^th^ dose of mRNA vaccines only restored the antibody titers to peak titers after the 3^rd^ dose, and the fold-increase of Omicron-NAbs was inferior to that of WT-NAbs (7.2 vs 11.4) after the 4^th^ dose of mRNA1273 ^11^. For BNT162b2, the neutralization breadth seemed to be narrowed too. Fold-difference of VNT50 between WT and Omicron was enlarged from 6.3 (after the 3^rd^ dose) to 11.3 (after the 4^th^ dose), despite that data came from live virus neutralization assays from two studies ^8,11^. These results are in line with our data.

Our results may provide several implications for the booster dose aiming to strengthen the protection against VOCs, such as Omicron. First, an urgent use of current inactivated vaccines for the 4^th^ booster is feasible but not ideal. Second, other types of vaccines may also suffer from RBD-NAbs suppression and shift of immune responses after repeated vaccination if the sequence keeps unchanged. Third, a recombinant S protein vaccine or mRNA vaccine based on the sequence of VOCs would be a good alternative for further boost, since mutated RBD may not be suppressed significantly by previous vaccination based on ancestral RBD. Even if that happened, mutated NTD capable of inducing NAbs against VOCs could compensate the loss of RBD-NAbs.

Our study has several limitations. First, the result came from a cohort of young HCWs. The effect of the 4^th^ dose on very young and elderly populations may be different. Second, only a pseudovirus neutralization assay was used in the current study ^13^. Nevertheless, our neutralization results regarding the ratio of NAb titers between WT and Omicron are in line with results from pseudovirus neutralization assay or authentic virus neutralization assay from other groups ^8,16,20^. Third, we did not assess other aspects of humoral immune responses such as antibody-dependent cellular cytotoxicity (ADCC) or the cellular arm of immunity. They may contribute to disease prevention even in the absence of NAbs.

In conclusion, our study demonstrated that the 4^th^ dose of inactivated SARS-CoV-2 vaccine is safe but the ability to further strengthen the protection against Omicron is compromised by suppression of RBD-NAbs and a further shift of humoral response away from RBD. Updated vaccines based on VOC sequences that take the advantage of RBD, NTD, and other antigenic domains would be an ideal alternative for future boosters.

## Methods

### Human subjects

In this study, we conducted a non-randomized trial and recruited participants from a prospective cohort at the First Affiliated Hospital of Sun Yat-sen University (FAH-SYSU) in Guangzhou, China. Sixty-three HCWs received a standard two-dose regimen of the inactivated SARS-CoV-2 vaccine (BBIBP-CorV, Sinopharm, Beijing) ^12^. Five months after the 2^nd^ dose, 50 of the 63 HCWs volunteered to receive a 3^rd^ dose of BBIBP-CorV ^2^. Thirty-eight of them volunteered for the current study aiming to investigate the safety and effectiveness of a 4^th^ dose, at the moment when healthcare systems were challenged by Omicron variant and the immune response induced by the 3^rd^ vaccination waned substantially. They received a 4^th^ homologous booster shot of the inactivated vaccine 6 months after the 3^rd^ vaccination. Blood samples were collected right before the booster dose, 14 days, and 28 days after the boost. All studies were approved by the Institutional Review Board of FAH-SYSU and written consent was obtained from all participants. The prospective cohort and the trial were registered to the Chinese Clinical Trial Registry (ChiCTR2100042222, ChiCTR2200055564).

### Blood samples

For serum collection, blood samples were allowed to clot at room temperature and subsequently centrifuged at 3000×g for 10 min. Sera were transferred into 0.5 ml aliquots in polypropylene tubes and stored at −80°C. To isolate peripheral blood mononuclear cells (PBMCs), blood samples were collected into the heparinized tubes. PBMCs were isolated by density-gradient centrifugation. Briefly, blood samples were diluted with PBS at a 1:1 ratio to 30 ml and loaded on top of 15 ml Lymphoprep™ (StemCell) in the 50 ml centrifugation tube and centrifuged at 800 rpm for 30 minutes. The medium cell layer was collected and washed with PBS once, followed by centrifugation at 400 rpm for 10 minutes. Pelleted PBMCs were cryopreserved in Bambanker (StemCell) immediately at -80°C.

### Cell lines and plasmids

Human ACE2 over-express HEK293T (hACE2-293T, PackGene Biotech) were cultured in DMEM (10-013-CVRC, Corning) supplemented with 10% fetal bovine serum (FBS, FSP500, ExCellBio), non-essential amino acids (NEAA, 11140-050, Gibco), 100 U/ml penicillin and 100 μg/ml streptomycin (SV30010, HyClone). Jurkat-Lucia™ NFAT-CD16 Cells (jktl-nfat-cd16, InvivoGen) was cultured in IMDM (BL312A, Biosharp) supplemented with 10% FBS, NEAA, 100 U/ml penicillin and 100 μg/ml streptomycin, 100 μg/ml Zeocin (ST-1450, Beyotime) and 10 μg/ml Blasticidin S (ST-018, Beyotime). All cell lines were passaged less than 15 generations and examined the mycoplasma by PCR and fluorescence labeling methods.

The plasmid pcDNA3.1-2019-nCoV-Spike is a gift from Dr. Lu Lu at Fudan University, encoding the spike protein from an ancestral SARS-CoV-2 reference strain (Wuhan-Hu-1) which is called as wild type (WT) throughout the manuscript. The plasmid pcDNA3.1(+)-Omicron-spike (JD20211214001R, Kidan Bio) encodes the spike protein from the Omicron variant (B1.1.529). The plasmid pcDNA3.1(+)-Envelope encodes envelope protein from WT was full-genome synthesized by Genewiz China according to the reference sequence NC_045512.2 in NCBI. Plasmids pSPAX2 and pLenti-CMV-Puro-Luc (168w-1) were a gift from Dr. Jianping Guo and purchased from MiaolingBio (P1216), respectively.

### ELISA

All SARS-CoV2 recombinant proteins were purchased from Sino Biological (Beijing, China). For ELISA, 200 ng/well of WT SARS-CoV2 spike (40589-V08B1), spike S1 subunit (40591-V08H), spike S2 subunit (40590-V08B), RBD (40592-V08H), nucleocapsid (40588-V08B) and envelope (40609-V10E3), Omicron spike (40589-V08H26), spike S1 subunit (40591-V08H41), RBD (40592-V08H121) and nucleocapsid (40588-V07E34) were coated on the 96-well ELISA plate (655061, Greiner Bio-one) using coating buffer (G3022, Saint Bio) overnight at 4 °C, respectively. Plates were washed by PBS supplemented with 0.5% Tween-20 (PBST) for three times, followed by blocking with 5% BSA in PBST (blocking buffer) for 1 hour at room temperature. Sera were firstly diluted 40-fold, followed by 4-fold serial dilution and incubation at 4°C overnight. Plates were washed 5 times by PBST, and incubated with 100 μl/well goat HRP conjugated anti-human IgG antibody (2040-05, SouthernBiotech, 1:3000) in PBST at room temperature for 30 min. After washing 5 times with PBST, 100 μl/well 3,3′,5,5′-Tetramethylbenzidine substrate (P0209, Beyotime) was added to each well for 15 min, stopped by the stopping buffer (P0215, Beyotime). OD450 was measured by Varioskan Lux Microplate Reader (Thermo Fisher).

### Pseudovirus neutralization assay

Pseudovirus production and neutralization assay were performed following a previous study ^13^. To generate WT SARS-CoV-2-Spike (Wuhan-Hu-1) pseudovirus, pcDNA3.1-2019-nCoV-Spike, pSPAX2 and pLenti-CMV-puro-Luc (168w-1) were co-transfected to HEK293T using Lipo8000 (C0533, Beyotime) according to the manufacturer’s instruction. For the generation of the B.1.1.529 Omicron-variant spike pseudovirus, pcDNA3.1(+)-Omicron-spike, pSPAX2 and pLenti-CMV-puro-Luc (168w-1) were co-transfected to HEK293T using Lipo8000. The virus-containing supernatant was harvested after 72 h and stored at −80°C until use. The hACE2-293T at 2×104/well were seeded on the black flat-bottom 96-well plate (655090, Greiner Bio-one) for 16 h in advance. Sera were firstly diluted 10-fold then 4-fold serial diluted subsequently in DMEM, then co-incubated with pseudovirus at 37°C for 1 h. The co-incubated samples, together with samples without sera or pseudovirus as controls, were subjected with 10 μg/ml polybrene (C0351, Beyotime) to the hACE2-293T for 6-hour absorption. The culture medium was replaced and incubated for another 42 h at 37°C. Infected cells were lysed by firefly luciferase lysis buffer (RG126M, Beyotime), then the luciferase substrate (RG058M, Beyotime) was applied for the luciferase assay according to the manufacturer’s instruction. The relative light unit (RLU) was measured by Varioskan Lux Microplate Reader (Thermo Fisher). The 50% pseudovirus neutralization titer (PVNT50) was determined by a four-parameter nonlinear regression curve (GraphPad Prism).

### Statistical analysis

Statistical analysis was performed using Graphpad Prism. Comparisons were assessed using Wilcoxon matched-pairs signed-rank test, paired Student’s t-test, Friedman test followed by Dunn’s multiple comparisons test, or RM One-way ANOVA as indicated. P values < 0.05 were considered as statistically significant.

## Data Availability

Authors will share de-identified individual participant data on request with researchers who provide a methodologically proposal and can conduct analyses that achieve the aims of the proposal. Reasonable requests for data sharing can be directed to by email. To gain access data requestors will need to sign a data access agreement.

## Acknowledgment

We thank Dr. Lu Lu at Fudan University for plasmids and kind help in pseudovirus neutralization assay. The work is supported by The Talent Program of the First Affiliated Hospital, Sun Yat-sen University (Y70311), The Hundred Talent Program of Sun Yat-sen University (Y61224), and the Science and Technology Program of Guangzhou (202103000076).

## Author Contributions

HX, SP and JW supervised the study. HX, SP and JW conceived and designed the study. YL, RM, CD and ML recruited participants in the trial and collected blood samples. CD, ML, YL, ZH, LS, JJ, LL and YLi performed the experiments and collected data. JW performed the statistical analysis. JW and CD drafted the manuscript, HX, SP, HZ and MinL made a critical revision. All authors approved the final version before submission.

## Conflict interests

The authors have no conflicts of interest to disclose.

## Figure Legends

**Supplementary Figure 1.**
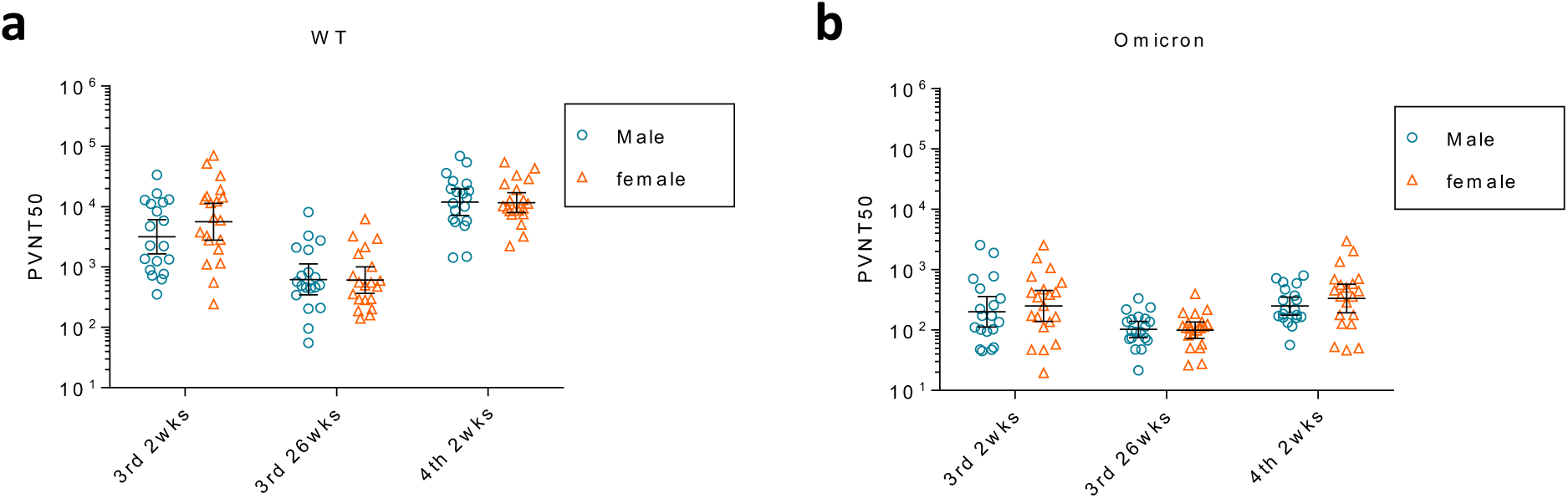
NAbs in males and females. Neutralization assays were performed to measure NAbs titers against pseudoviruses with S protein from a WT strain or Omicron variant. (**a**) NAb titers for WT virus in males (blue circle) or females (orange triangle) were compared. (**b**) NAb titers for Omicron variant were similarly compared between males (blue circle) or females (orange triangle). Data were shown as Geometric mean ± 95% Cl.

**Supplementary Figure 2.**
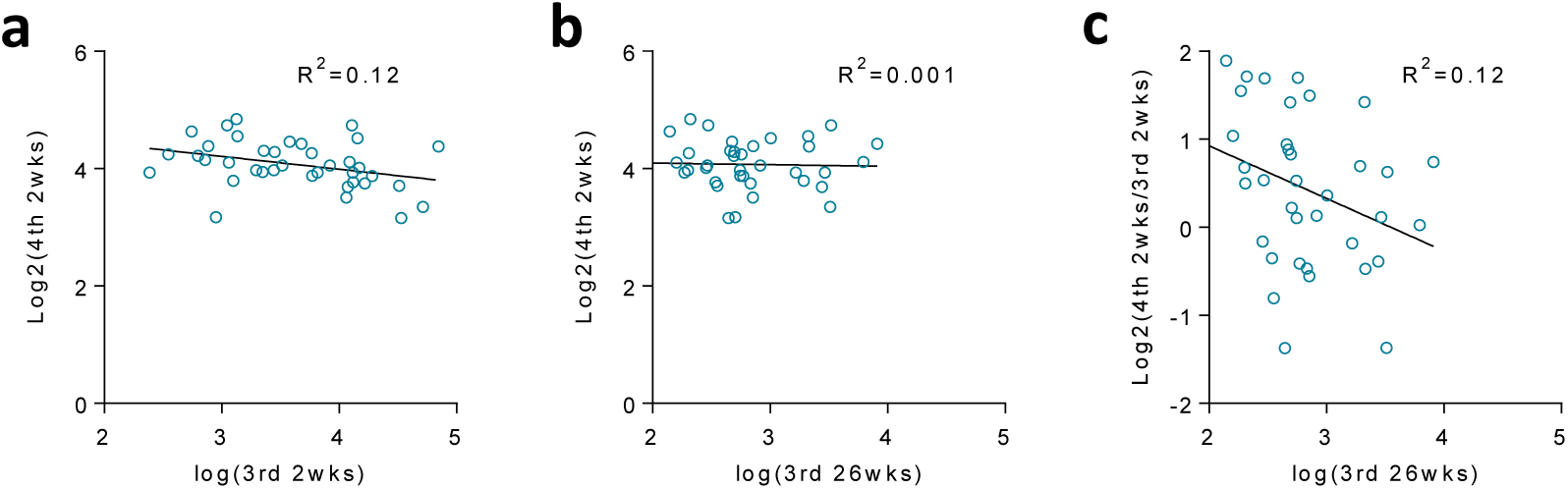
Correlation between NAb levels at various time points. Linear regressions were performed to reveal the potential correlation between NAb levels at 3^rd^ 2wks and 4^th^ 2wks (**a**), or NAb levels between 3^rd^ 26wks and 4^th^ 2wks (**b**), or fold change of peak values and NAb level at 3^rd^ 26wks (**c**).

**Supplementary Figure 3.**
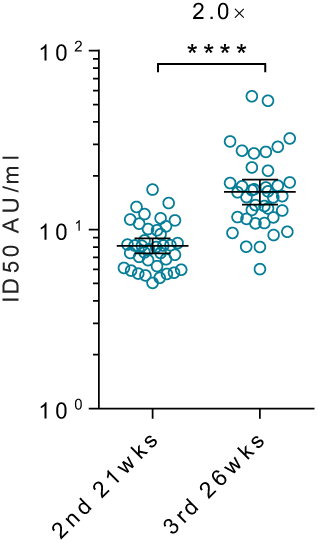
Comparison of RBD-NAbs at 2nd 21wks and 3rd 26wks. RBD-NAbs were measured by a one-step competitive Chemiluminescent immunoassay. RBD-NAbs at 2^nd^ 21wks and 3^rd^ 26wks were compared. Data were shown as Geometric mean ± 95% Cl. Paired t-test was used. ****p<0.0001.

**Supplementary Figure 4.**
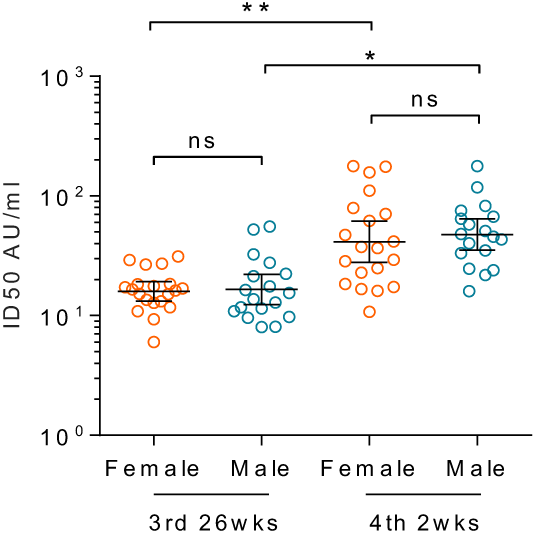
RBD-NAbs in males and females. RBD-NAbs were measured by a one-step competitive Chemiluminescent immunoassay. RBD-NAb titers in males (blue circle) or females (orange triangle) at 3^rd^ 26wks or 4^th^ 2wks were compared. Data were shown as Geomean±95% CI. Data were shown as Geometric mean ± 95% Cl. RM one-way ANOVA was used. * p<0.05, **p<0.01. ns, not significant.

## Notes

### Competing Interest Statement

The authors have declared no competing interest.

### Clinical Trial

ChiCTR2200055564

### Author Declarations

The Institutional Review Board of FAH-SYSU gave ethical approval for this work.

